# An mHealth intervention based on behavior change techniques to promote physical activity and nutrition in older patients with cancer: protocol for an N-of-1 trial

**DOI:** 10.64898/2026.07.06.26356658

**Authors:** Mathis Brusseau, Julie Deffrennes, Blandine Gallet-Suchet, Laurence Cristol, Gérard Dray, Sophie Gendrault, Lobna Harguem, Rémy Dadier, Julie Boiché

## Abstract

**BACKGROUND:** Older adults with cancer often struggle to achieve recommended levels of physical activity and dietary intake. Ecological momentary assessment combined with accelerometry can provide insights into the temporal dynamics of psychological and behavioral processes at the individual level, such as motivation towards health behaviors.

**OBJECTIVE:** This N-of-1 study aims to improve physical activity and dietary intake among older patients with cancer using an mHealth behavioral intervention.

**METHODS:** A single-subject ABA’ design will be employed among older patients with cancer (≥ 70 years). A 2-week baseline phase (A) will be followed by an 8-week intervention phase (B) and a 2-week follow-up phase (A’). Throughout all these phases, participants will complete a daily data collection protocol combining ecological momentary assessment questionnaires and an ActiGraph wGT3X-BT accelerometer worn on the waist to measure physical activity. Ecological momentary assessment questionnaires will be delivered via a digital application to collect information on dietary intake, fatigue, and motivational constructs based on the Theory of Planned Behavior. The intervention (B) will consist of an mHealth intervention based on behavior change techniques, delivered via weekly calls, personalized messages, and a digital application. Data will be analyzed using piecewise regression models. In addition, a semi-structured interview will be conducted to assess patient experience. These qualitative data will help identify contextual factors, such as treatment-related side effects or variations in health status, that may have influenced behavior change and participation in data collection.

**CONCLUSION:** This N-of-1 study explores intra-individual behavioral dynamics using intensive longitudinal data rather than testing a strictly reversible intervention effect. The mHealth intervention is based on behavior change techniques, with adjustments tailored to participants’ progress and changing circumstances. The results will contribute to the evidence base for mHealth interventions designed to promote sustained physical activity and dietary intake among older adults with cancer.

**Trial registration:** ClinicalTrials.gov (NCT06445140), registered on 06 June 2024.

## Introduction

Cancer is predominantly an age-related disease. As the global population continues to age, the number of older adults living with cancer is expected to rise significantly worldwide [1], especially among patients aged over 80 years [2]. This population is underrepresented in clinical trials [3,4], especially those focusing on physical activity [5] and mHealth interventions [6]. The risk of engaging in unhealthy behaviors, such as physical inactivity and malnutrition, increases with age and becomes even more pronounced during cancer treatment [7].

Reduced food intake is a well-established cause of malnutrition, which remains a significant concern among older patients with cancer [8]. Similarly, physical inactivity is widespread and problematic among older people, particularly in geriatric oncology [7,9]. These put patients at risk of poor health outcomes and contribute to the functional decline observed in older patients 2–3 months after the start of chemotherapy [10]. Therefore, the inclusion of dietary components within supportive care interventions [11,12] and exercise interventions is essential and increasingly recognized among this population [13,14]. Tailored exercise plans, behavioral strategies, and nutritional support could mitigate treatment side effects, such as those associated with chemotherapy agents, and promote healthier and more effective ageing trajectories [15]. With this aim, behavior change interventions and adherence support are required to help older adults increase their physical activity levels and improve dietary intake in daily life, especially during cancer treatment [16]. A meta-analysis has shown that simultaneous modifications of multiple health behaviors are possible to manage chronic conditions among patients with an average age of 60 [17]. Health behaviors do not necessarily operate independently but often co-occur [18]. Concepts such as behavioral spillover, transfer, and compensation provide potential frameworks for understanding these interrelationships. Whether such processes occur between physical activity and dietary intake in older adults with cancer remains to be investigated. In geriatric oncology, behavioral interventions targeting nutrition and physical activity simultaneously are therefore expected to be effective.

Furthermore, the use of theoretical frameworks may help to better understand health behaviors in older adults [19,20]. Although theory-based approaches have shown promising results, further research is needed, particularly among older adults with cancer. Specifically, the theory of planned behavior (TPB) has been widely used in previous studies on physical activity and nutrition [20–24]. According to the TPB, behavior emerges from individual’s intentions, which are influenced by perceived behavioral control, instrumental and affective attitudes, and subjective norms. Furthermore, interventions targeting TPB variables appear to be effective in increasing physical activity and dietary intake in cancer patients and geriatrics [25,26]. Physical activity and dietary behaviors may be influenced by changes in perceived benefits, social support and perceived ability to engage in these behaviors in the context of treatment-related challenges. TPB therefore provides a relevant framework for identifying modifiable motivational factors that can be targeted through tailored behavioral interventions.

To this end, collecting daily data is necessary to gain detailed insights into the real-time dynamics of health behavior [27–29]. As health status can fluctuate, especially during treatment, patients’ engagement in supportive care interventions needs to be assessed and addressed [30]. The combination of wearable technology and Ecological Momentary Assessment (EMA) is an innovative and feasible method for collecting behavioral data [31] and for monitoring within-person dynamic processes in real-world settings. These two methods achieve high compliance levels among older patients with cancer [32,33]. Thus, mHealth interventions are particularly relevant for evaluating and delivering behavioral interventions by providing repeated cues and measurements [34], especially in at-risk and medically vulnerable populations [35,36]. In this context, regular phone calls are effective among older patients with cancer [30] and can be adapted over time to reflect changes in health status or capabilities during treatment, alongside personalized messages [37]. Notably, 85% of patients with cancer have expressed a desire to receive weekly phone calls during chemotherapy to support exercise adherence [38].

Among the components of behavioral interventions, behavior change techniques (BCTs) offer a promising approach to increase physical activity and nutrition in cancer patients and survivors [22,27,39–41]. BCTs are effective in strengthening intentions and perceived behavioral control related to physical activity and nutrition [22,42]. More specifically, in behavioral interventions with activity trackers targeting older adults, the most commonly used BCTs are goal setting, self-monitoring, and feedback on behavior [27,43]. These strategies may contribute to greater awareness and motivation among participants by offering clear targets and sustained reinforcement, which could in turn facilitate self-regulation. However, it is well-established that all intentions do not always translate into actual behavior, a phenomenon known as the intention-behavior gap [44]. To bridge this gap in physical activity, specific BCTs offer promising strategies to translate intentions into action effectively [45]. Key approaches include enhancing self-regulation (e.g., action planning, self-monitoring), strengthening perceived behavioral control (e.g., problem-solving, coping planning), and promoting habit formation (e.g., cue-based repetition) [45].

In the context of geriatric oncology, behavioral intervention effects can be difficult to detect using traditional pre- and post-intervention measurements. These challenges are partly explained by time-varying influences, such as cancer treatment effects and fluctuations in health status in older adults, which may bias the estimation of intervention effects. Thus, a useful method to examine the evolution of behaviors and motivation during intervention and non-intervention phases is the single-case experimental study (N-of-1). Single-case experimental designs rely on repeated measurements over time, allowing evaluation of intervention effects at the individual level. This is particularly relevant for behavioral interventions characterized by high interindividual variability and context-sensitive responses [46,47]. The N-of-1 design has attracted growing interest in oncology [37,48], physical activity [49,50], and nutrition research [51,52]. Previous studies have used N-of-1 designs to investigate behavior change techniques in cancer populations [48], while other studies have combined N-of-1 designs with EMA to examine daily changes in physical activity [49,53]. This approach has also been applied in women with breast cancer to examine daily changes in psychological outcomes during a physical activity intervention [37]. However, the potential of personalized, theory-informed mHealth interventions integrating behavior change techniques to promote physical activity and dietary intake in older adults with cancer remains underexplored.

## Materials and methods

This protocol is the interventional part of the MONAGE project: a single-case experimental design with an ABA’ structure employed among older patients with cancer (≥ 70 years). A 2-week baseline phase (A) will be followed by an 8-week intervention phase (B) and a two-week follow-up phase (A’). Throughout these three phases (A-B-A’), participants will engage in a daily data collection protocol combining EMA and the use of a wGT3X-BT accelerometer (ActiGraph, Pensacola, FL, USA). The intervention, which will be delivered only during phase B, will consist of an mHealth intervention based on BCTs, carried out via weekly calls and personalized messages throughout the week. This manuscript was written according to the Single-Case Reporting Guideline in BEhavioral Interventions (SCRIBE) [54]. The SCRIBE checklist is provided in the supplementary materials (S1).

The primary objective of the interventional part of MONAGE is to increase the daily duration of moderate-intensity physical activity, measured using the ActiGraph wGT3X-BT accelerometer, and dietary intake, assessed using a daily 10-point self-reported numeric rating scale, following an mHealth intervention in older patients with cancer.

We hypothesized that the intervention will increase the level of physical activity and dietary intake, thanks to the changes in motivational factors targeted by the intervention, namely intentions and perceived behavioral control. These changes were expected to occur during the intervention phase (B) compared with baseline (A) and to be slightly attenuated during the follow-up phase (A’).

### Patient selection

The aim is to complete recruitment by December 2026, enrolling a total of 15 patients. Given the N-of-1 design, the study primarily focuses on within-person changes and repeated measurements rather than on a large sample size for between-group comparisons [55]. Patients attending the Montpellier Cancer Institute (ICM) for oncogeriatric care will be considered for inclusion and invited to participate by the investigator. Patients must meet the criteria previously described in the observational phase of MONAGE [56], and the recruitment process follows the same procedure. Briefly, patients aged ≥ 70 years, diagnosed with a solid tumor located at any site, with a Geriatric 8 (G8) score ≤ 14, and with a moderate-intensity physical activity level <150 minutes/week, who provided informed and written consent, will be included. Patients with brain metastases; those unable to use the digital platform or perform physical tests; those who are unable to eat orally; those with contraindication or inability to perform physical activity; those unable to attend regular follow-up due to psychological, family, numerical, social or geographical reasons; and those lacking legal capacity or under protective custody or guardianship will not be included. Written informed consent will be obtained from all participants prior to inclusion in the study.

### Procedures

#### Baseline assessments and consultations

Each patient will attend three consultations: one with a geriatric oncologist, one with a kinesiologist specialized in adapted physical activity, and one with a dietician. A detailed description of the content of these consultations has been provided previously [56]. The patient will undergo a Comprehensive Geriatric Assessment (CGA) with a geriatric oncologist, including the completion of the Geriatric Core Data Set (G-CODE) [57]. During the exercise consultation, the patient will receive the *Institut national du Cancer* (INCa) recommendations for physical activity [58], which include: ≥150 minutes of moderate-intensity cardiorespiratory exercise per week, 2 sessions of muscle-strengthening exercise per week, 2 to 3 sessions of flexibility and mobility exercise per week and balance exercises twice a week for patients aged ≥65. During the nutritional consultation, the patient will receive three recommendations: incorporate protein and energy-rich foods into the diet; eat small and frequent meals to ensure adequate intake; and drink water between meals to maintain appetite. If necessary, patients will be prescribed physiotherapy and oral nutritional supplements.

Assessments conducted during the exercise and nutritional consultations will include: physical activity level (Global Physical Activity Questionnaire; [59]), grip strength (Handgrip with Jamar PLUS+ Digital Dynamometer; [60]), lower extremity function (Short Physical Performance Battery; [61]), dietary intake (Visual/Verbal Analogue Scale of food ingesta (ingesta-VVAS); [8,62]) and nutritional risk (Mini Nutritional Assessments; [63]).

These initial consultations may already involve several BCTs, including feedback on behavior, instruction on how to perform the behavior, information about health consequences, and credible source. These components are part of usual clinical care and therefore represent potential pre-intervention exposure before the start of the mHealth intervention.

#### Study design

After completion of baseline assessments and consultations, a single-case experimental design (N-of-1, ABA′) will be implemented. During phase A (baseline), motivation, fatigue and target behaviors will be measured repeatedly. This phase will last 2 weeks, a duration supported by the feasibility and adherence observed during the initial observational phase of the MONAGE research project. This phase A will serve to establish a baseline observation phase prior to the delivery of the mHealth intervention. During phase B, an experimental behavioral intervention will be introduced. This intervention will consist of an 8-week mHealth intervention based on BCTs, delivered through one weekly telephone call, personalized messages, and the use of a digital application. Throughout phase B, target variables will continue to be assessed daily. During phase A’, the intervention will be withdrawn while measurement of target behaviors will continue using the same procedures as in the baseline phase. This phase will last two weeks and will aim to assess whether changes observed during the intervention phase are maintained. Figure 1 presents the overall study design, including the initial visit, the ABA’ structure, and the end-of-study visit. Medical consultations and supportive care may be provided as part of the usual care, if needed, during the study.

**Figure 1:**
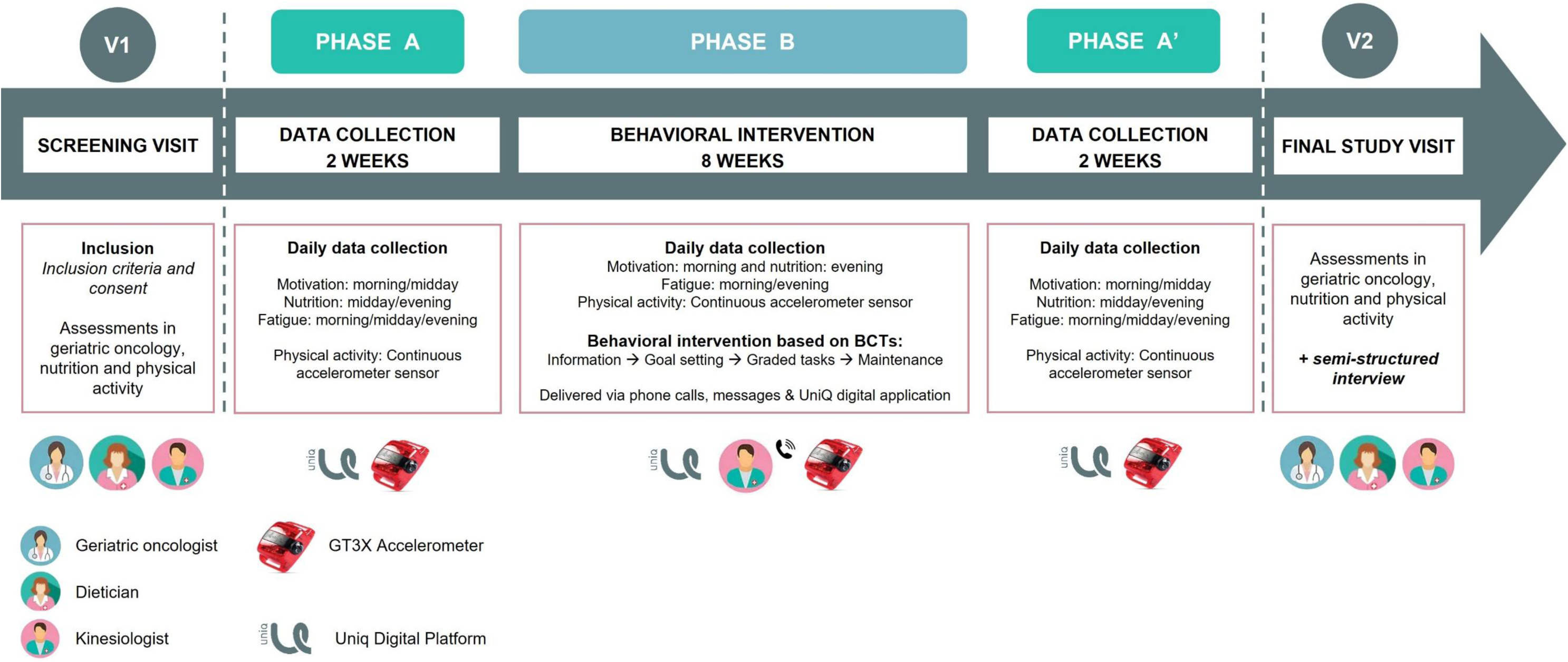
Overview of the ABA’ design and assessments

#### Final study visit: assessments and semi-structured interview

At the final study visit, patients will repeat the three initial consultations and will participate in a semi-structured interview. Conducting a semi-structured interview at the end of an N-of-1 behavioral intervention allows for an in-depth exploration of the participant’s experiences [64]. These interviews help identify contextual factors, such as treatment-related effects or variations in health status, that may have influenced the intervention outcomes. While N-of-1 designs provide detailed longitudinal quantitative data at the individual level, they may not fully capture the mechanisms underlying observed changes in behavior and health status. The interview guide, translated into English, is presented in Table 1. Items in parentheses represent optional probing prompts used by the interviewer, when appropriate, to encourage participants to elaborate on aspects not spontaneously addressed.

**Table 1.** Questions planned for the semi-structured interview after completion of the entire ABA’ design.

| Themes | Questions |
| --- | --- |
| Adherence to the intervention | Q1.1: You took part in a 12-week experiment involving monitoring your health behaviors and interventions such as phone calls and messages. How would you describe your overall experience?<br>Q1.2: What did you think of the phone calls and messages with the professional in charge of this intervention?<br>Q1.3: Did you find the questionnaires and the accelerometer easy to use? |
| Behavioral effects of the intervention | Q2.1: Could you explain what physical activities you've been doing? How have these activities changed over the course of the intervention? (*e.g. frequency, intensity, duration, type*)<br>Q2.2: Could you describe your eating habits? Did they change at any point during the intervention? |
| Impact of external constraints | Q3.1: Aside from the intervention, what factors have influenced your physical activity and eating habits over the past few weeks? (*e.g. side effects of treatment, support from relatives*) |
| Motivational effects of the intervention | Q4.1: Has your motivation changed since this support program was implemented?<br>Q4.2: What role did this intervention play in achieving your physical activity and nutrition goals? |
| Feedback and perspectives | Q.5.1: What improvements would you suggest to make this intervention better meet your expectations?<br>Q.5.2: Is there anything else you would like to add? |

### Outcomes measures

In phase A, data collection will begin on the day following inclusion. Assessments of patients during phases A and A’ will serve as baseline and post-intervention evaluations, respectively. Data collected will be the same as those collected during the 2-week observational phase of MONAGE [56]: motivation based on the Theory of Planned Behavior (morning and midday), nutrition (midday and evening), and fatigue (morning, midday and evening). All assessment items are provided in supplementary material (S2). Data will be collected via questionnaires on the UniQ application using an EMA approach, as well as with a wGT3X-BT accelerometer worn on the waist. We will test the UniQ application with each participant before the study begins to ensure they are comfortable using it. The application has a simple and user-friendly interface, as illustrated in supplementary materials (S3).

During phase B, which lasts eight weeks, data collection will be simplified by removing the midday questionnaire. It will include a morning questionnaire assessing motivation and fatigue, and an evening questionnaire assessing self-report behaviors and fatigue, presented in the supplementary materials (S4).

Physical activity data will be measured using an accelerometer (ActiGraph wGT3X-BT), which provides a reliable, valid, and direct measure [65]. In line with current recommendations for accelerometry reporting, detailed information on device characteristics is provided [66]. Participants will be instructed to wear the device on the waist all day for the entire data collection period, except during sleep. The accelerometer will be worn throughout the ABA’ design. A valid day will be defined as a day during which the participant wears the device for at least 8 hours [67]. Raw triaxial acceleration data will be collected at a sampling frequency of 30 Hz. Physical activity will be derived from raw acceleration data using the Euclidean Norm Minus One (ENMO), expressed in milligravity (mg). The objective is to identify intensity thresholds based on ENMO to distinguish inactivity from light, light from moderate physical activity, respectively. Based on studies involving an elderly population, we will use the following cutoff points: 14 for light (threshold.lig) and 70 for moderate (threshold.mod) intensity physical activity [68,69]. The M30 and M60 metrics will be calculated from ENMO values (mg), representing the minimum acceleration during the most active 30 and 60 minutes of the day, respectively. Data from ActiGraph wGT3X-BT will be analyzed using the GGIR R-package [69].

### Intervention

The behavioral intervention has been approved and revised by patients, patient partners, geriatric oncologists, dieticians, adapted physical activity teachers (kinesiologists), health project managers, and behavior change specialists. More precisely, the assessment items were co-designed and validated with patients. The intervention was refined through one or two individual consultations with the relevant professionals to gather feedback, followed by one group consultation for discussion and validation with the geriatric oncologists.

This intervention will consist of delivering an mHealth behavioral intervention based on BCTs over 8 weeks, involving one telephone call per week and personalized messages. The BCTs included in the intervention were selected primarily to target intentions and perceived behavioral control, as these were identified as relevant motivational factors associated with significant changes in physical activity and dietary intake during the observational phase of MONAGE. Thus, the observational findings directly informed the selection of the TPB constructs targeted in the intervention.

For this purpose, BCTs were selected based on the Behavior Change Technique Ontology (BCTO) [70] and evidence from the Theory and Techniques Tool, which links BCTs to their proposed mechanisms of action [71]. The primary mode of delivery for BCTs will be individual-based and delivered through phone calls. A secondary mode of delivery will be provided via SMS messages and the use of a mobile application for daily activity reporting.

The predefined set of BCTs used during the weekly calls and the intervention objectives are presented in Table 2. The intervention is structured into four components: content familiarization (B1), goal setting (B2), learning and graded tasks (B3), and habit reinforcement (B4). For component B1, BCTs similar to those potentially delivered during the initial consultations will be implemented but will be informed by participants’ behaviors observed during Phase A. Identified as core BCTs of the intervention, Goal setting, Action planning, Self-monitoring, and Goal review form a recurring cycle throughout the intervention, allowing goals and action plans to be revisited and adjusted according to participants’ progress and changing circumstances. This flexible sequencing is consistent with recent work suggesting that BCTs may be selected and revisited according to participants’ current phase of behavior change, relevant determinants, and emerging barriers [72].

**Table 2.**
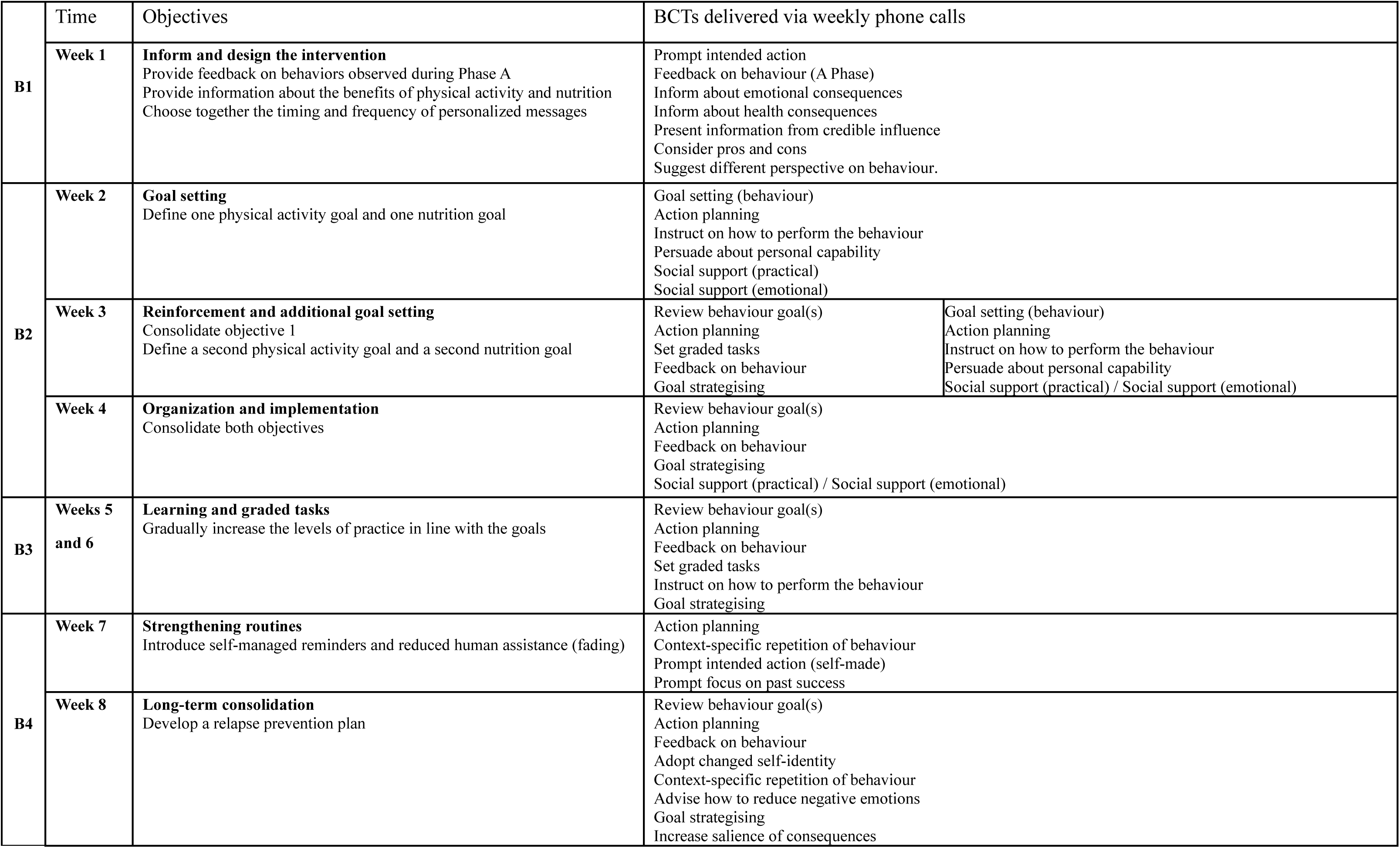
Description of the intervention, including its objectives and associated behavior change techniques during the weekly phone calls.

In addition, two BCTs were predefined as adaptive components: Monitoring of emotional consequences and Manage negative emotions. Their use may be informed by weekly discussions with the participants, treatment-related changes, or changes in health status. These techniques were selected because emotional responses may vary substantially across participants and over time, particularly in the context of cancer treatment. If used, adaptive BCTs will be documented for each participant.

To assess intervention fidelity, telephone calls will be audio-recorded and independently reviewed. Intervention fidelity could therefore be considered at two complementary levels: delivery fidelity, referring to whether the predefined BCTs were delivered as intended, and adaptation fidelity, referring to whether adaptive BCTs were selected and delivered.

To promote physical activity while minimizing environmental and financial barriers, the objectives will focus on accessible activities such as walking, as well as independent or supervised muscle-strengthening exercises. For nutrition, the objectives will focus on increasing daily calories and protein intake and/or ensuring adherence to prescribed oral nutritional supplements. Throughout the intervention, patients will report their behaviors daily using the digital application (self-monitor behaviour BCT), presented in supplementary material (S4). During the week, personalized messages will be sent at a frequency determined by the patient. The proposed schedule includes a message on day three following the call to reiterate the objectives (Prompt intended action BCT), followed by a message on day five providing feedback based on participants’ reported daily behaviors (Feedback on behaviour BCT). For example, a personalized message sent on day five could be: “I have reviewed your questionnaire responses. You were able to go for a walk every other day, and you managed to take your oral nutritional supplement every day. Well done on your efforts this week! You can keep this up until our call on Wednesday.”. From week 5 onwards, the frequency of these messages may be reduced or discontinued to progressively limit external support.

### Safety monitoring

Participants’ ability to engage in physical activity will be assessed during the initial consultations, and physical activity recommendations will be adapted to their functional capacity and health status. When independent practice is not appropriate, supervised care, including physiotherapy, will be recommended. Participants will also be instructed to report any exercise-related symptoms, adverse events, or falls occurring during the intervention, which could lead to adaptation or interruption of the recommended activities.

### Statistical analysis

#### Quantitative analysis

We will evaluate the participants’ adherence to and acceptance of the data collection using descriptive statistics, including means, medians, and percentages. These measures will be used to summarize the key characteristics of the dataset. Each participant’s data will be treated as a distinct dataset and analyzed individually. The primary sample size consideration was the number of repeated observations per participant. The planned repeated daily assessments provide up to 86 observations (15 in A, 56 in B, and 15 in A′) per primary outcome and participant, allowing detailed estimation of within-person trajectories [55]. Missing data will be handled within a maximum likelihood framework under the assumption of missing at random (MAR), which will be assessed through patterns of missingness over time and in relation to observed outcomes. Piecewise regression models will be fitted using all available observations without prior smoothing. Sensitivity analyses may explore state-space modeling approaches (e.g., Kalman filtering) to account for intermittent missingness.

As a first step, daily trajectories of physical activity (duration at various intensity levels, M30, ENMO), dietary intake (Visual/Verbal Analogue Scale of food ingesta), motivation and fatigue will be plotted for each participant across the study period and visually inspected for trends and anomalies, considering the treatments.

The primary physical activity outcome will be daily moderate-intensity physical activity duration (minutes/day), while other accelerometry-derived metrics will be considered as secondary or exploratory outcomes. The primary nutritional outcome will be dietary intake measured each evening using the daily 10-point numeric rating scale based on the ingesta-VAS. Changes in level and slope across phases A, B, and A′ will be estimated using piecewise linear regression, separately for moderate-intensity physical activity duration and dietary intake, following the approach described by Wilbert in 2026 and implemented using the scan package in R [73]. Serial correlation between repeated daily observations will also be assessed and accounted for in the regression models where appropriate.

### Qualitative analysis

Semi-structured interviews will be conducted by two researchers independent of the intervention, using the interview guide, and audio-recorded using a mobile phone. Recordings will be transcribed using Microsoft Word’s transcription tool and checked while carefully proofreading the text. Two researchers, independent of the intervention, will code the transcripts using QualCoder. Following Braun and Clarkes’ approach, an inductive thematic analysis will be carried out in six phases, during which researchers may have to go back and forth [74]. The six phases include: familiarization, code generation, theme and category construction, revision, redefinition, and final report production. Thus, codes and themes will subsequently be compared, discussed, and refined through consensus with a third researcher who will not be involved in the coding, providing an external perspective on the interpretation. Data saturation using the method described by Guest et al. will be performed [75]. To evaluate the quality and rigor of this qualitative research, Braun et al.’s 15-point checklist of criteria for good thematic analysis will be applied [74].

## Discussion

### Expected findings

This study aims to improve physical activity and nutritional behaviors among older patients with cancer through an mHealth behavioral intervention incorporating BCTs. First, it is expected that patient participation results in a high completion rate for the EMA assessments [33] and ActiGraph wGT3X-BT wear time [32], around 70%. Similarly, we expect strong adherence to the behavioral intervention regarding phone calls [38]. Older adults with cancer are more likely to participate in cancer research when trial designs are adapted to their needs and feasible within their health and care contexts [4].

This study is designed to closely reflect real-world conditions experienced by older patients with cancer undergoing treatment. Physical activity objectives will focus on accessible activities such as walking, as well as independent or supervised muscle-strengthening exercises. The medical team prescribes physical therapy sessions or oral nutritional supplements if they determine that these interventions are clinically indicated. The overall aim is to promote physical activity and dietary intake under optimal conditions for patients.

### Limitations and strengths

Several limitations of the study should be considered. First, the eligibility criteria do not allow full representation of the older cancer population. The recruitment process may lead to selection bias, as participants who are more motivated, functionally able, or comfortable with technology may be more likely to participate. For example, active patients, individuals with low digital literacy, and non-users of digital technologies are excluded. In addition, the sample size does not allow generalization to the entire patient population, especially given its important heterogeneity. A wide range of sociodemographic profiles and cancer diagnoses cannot be fully represented. Cancer type and treatment characteristics may contribute to differences in physical activity levels, dietary behaviors, and intervention adherence; however, the N-of-1 design precludes meaningful comparisons between cancer types. Furthermore, this study cannot be conducted or continued if a patient requires major surgery. Because behavioral interventions often have cumulative and long-lasting effects and are influenced by external factors such as treatment side effects and changes in health status, the return-to-baseline phase (A’) may not fully reflect the pre-intervention state. This limits the ability to interpret reversibility within the ABA’ design.

A key strength of this study is the implementation of a structured intervention in a real-world clinical setting that accounts for changes in health status during treatment. The intensive longitudinal assessment, combining ActiGraph wGT3X-BT and EMA, further enables the detection of short-term behavioral fluctuations and individual-level intervention effects. Finally, the combination of an N-of-1 design with a digitally delivered, personalized BCT intervention, enables behavior change strategies to be tailored to individual needs and responses over time [76]. Moreover, patients will undergo a complete assessment at baseline and post-intervention using the Comprehensive Geriatric Assessment tool, along with specific evaluations of physical activity and nutrition. Qualitative analysis will help contextualize the effects of the intervention. The team developed this intervention protocol collaboratively, including health professionals and behavior change specialists. Quantitative and qualitative findings will be analyzed separately; however, their subsequent consideration together will provide complementary perspectives on the intervention and enable a context-specific interpretation of its impact, particularly by contextualizing individual quantitative trajectories.

### Future directions

This N-of-1 ABA’ study explores intra-individual behavioral dynamics using intensive longitudinal data rather than testing a strictly reversible intervention effect. Examining the relationship between motivation and behaviors in real time may yield valuable insights into the psychological processes underlying physical activity and nutrition in older adults with cancer. This project will provide insights into how behavioral patterns evolve during treatment and how these dynamics interact with fluctuations in fatigue and motivation. Furthermore, investigating the effects of an mHealth intervention grounded in BCTs on daily and within-day behavioral patterns represents a promising direction for advancing behavior change in this underrepresented population. The results of this study will inform the development of future individualized interventions aimed at improving adherence to physical activity and nutritional interventions. In summary, this study seeks to elucidate the mechanisms linking motivation and behaviors to optimize care pathways. If replicated in larger studies, this mHealth-based behavioral intervention may have potential for broader clinical use in this population.

## Abbreviations

BCT: Behavior change technique
BCTO: Behaviour Change Technique Ontology
CGA: Comprehensive Geriatric Assessment
EMA: Ecological Momentary Assessment
G8: Geriatric 8
G-CODE: Geriatric Core Data set
ICM: Montpellier Cancer Institute
INCa: Institut National du Cancer
SCRIBE: Single-Case Reporting Guideline in BEhavioural Interventions
TPB: Theory of Planned Behavior

## Declarations

### Ethics approval and consent to participate

This research project has been approved by the national ethics committee (Comité des Personnes Nord Ouest III, accepted on 30 May 2024) and the ANSM (French National Agency Authority for the Safety of Health Products) has been informed (19 June 2024). This research will be conducted in accordance with the French Public Health Code, the Good Clinical Practice and the Declaration of Helsinki. This study has been registered on clinicaltrial.gov (NCT06445140), 06 June 2024.

## Data Availability

No datasets were generated or analyzed during the current study.

## Acknowledgements

We thank Dr Fanny Salasc at the Montpellier Cancer Institute, France, for providing medical writing and editorial support. We would like to sincerely thank the patients and the healthcare professionals from Move in Med, Montpellier Cancer Institute, and EuroMov Digital Health in Motion for their valuable contributions to the refinement and validation of the intervention through their participation in its review and improvement.

A preprint version of this article has been peer-reviewed and recommended by PCI Health & Mov Sci (https://doi.org/10.24072/pci.healthmovsci.100542).

## Consent for publication

Not applicable

## Availability of data and materials

No datasets were generated or analyzed during the current study.

## CRediT authorship contribution statement

**Mathis Brusseau:** Conceptualization, Methodology, Software, Investigation, Writing – original draft, Visualization, Project administration, Funding acquisition. **Julie Deffrennes:** Conceptualization, Methodology, Project administration, Supervision, Writing – review & editing. **Blandine Gallet-Suchet:** Conceptualization, Funding acquisition, Writing – review & editing. **Laurence Cristol:** Methodology, Writing – review & editing. **Gérard Dray:** Methodology, Writing – review & editing. **Sophie Gendrault:** Methodology, Resources, Writing – review & editing. **Lobna Harguem:** Project administration, Writing – review & editing. **Rémy Dadier:** Methodology, Visualization, Writing – review & editing. **Julie Boiché:** Conceptualization, Methodology, Supervision, Writing – review & editing.

