## Supplementary files for "An mHealth intervention based on behavior change techniques to promote physical activity and nutrition in older patients with cancer: protocol for an N-of-1 trial"

**SUPPLEMENTARY MATERIALS**

- S1: The Single-Case Reporting Guideline In BEhavioural Interventions (SCRIBE) 2016 Checklist
- S2: EMA assessment items: The 5 versions of the questionnaire for TPB constructs, dietary intake and fatigue
- S3: Visual representation of the display of UniQ application
- S4: Self-monitoring of behaviors conducted on the UniQ application
- **S1: The Single-Case Reporting Guideline In BEhavioural Interventions (SCRIBE) 2016 Checklist**

|  | Item Number | Item description | Validation |
| --- | --- | --- | --- |
| **TITLE and ABSTRACT** | 1 | Identify the research as a single-case experimental design in the title | Yes |
|  | 2 | Summarize the research question, population, design, methods including interventions (independent variable/s) and target behavior/s and any other outcome/s (dependent variable/s), results, and conclusions | Yes |
| INTRODUCTION | | |  |
| Scientific background | 3 | Describe the scientific background to identify issues under analysis, current scientific knowledge, and gaps in that knowledge base | Yes |
| Aims | 4 | State the purpose/aims of the study, research question/s, and, if applicable, hypotheses | Yes |
| METHOD | | |  |
| Design | 5 | Identify the design (e.g., withdrawal/reversal, multiple-baseline, alternating-treatments, changing-criterion, some combination thereof, or adaptive design) and describe the phases and phase sequence (whether determined a priori or data-driven) and, if applicable, criteria for phase change | Yes |
| Procedural changes | 6 | Describe any procedural changes that occurred during the course of the investigation after the start of the study | Yes |
| Replication | 7 | Describe any planned replication | No formal replication across participants or conditions was planned, as this is a single-case ABA’ design. |
| Randomization | 8 | State whether randomization was used, and if so, describe the randomization method and the elements of the study that were randomized | Not performed, as the study uses a fixed ABA’ single-case experimental design. |
| Blinding | 9 | State whether blinding/masking was used, and if so, describe who was blinded/masked | Blinding was not implemented due to the nature of the behavioral intervention and outcome assessment. |
| PARTICIPANTS |  |  |  |
| Selection criteria | 10 | State the inclusion and exclusion criteria, if applicable, and the method of recruitment | Yes |
| Participants characteristics | 11 | For each participant, describe the demographic characteristics and clinical (or other) features relevant to the research question, such that anonymity is ensured | Yes |
| CONTEXT |  |  |  |
| Setting | 12 | Describe characteristics of the setting and location where the study was conducted | Yes |
| APPROVALS |  |  |  |
| Ethics | 13 | State whether ethics approval was obtained and indicate if and how informed consent and/or assent were obtained | Yes |
| MEASURES and MATERIALS |  |  |  |
| Measures | 14 | Operationally define all target behaviors and outcome measures, describe reliability and validity, state how they were selected, and how and when they were measured | Yes |
| Equipment | 15 | Clearly describe any equipment and/or materials (e.g., technological aids, biofeedback, computer programs, intervention manuals or other material resources) used to measure target behavior/s and other outcome/s or deliver the interventions | Yes |
| INTERVENTIONS |  |  |  |
| Intervention | 16 | Describe the intervention and control condition in each phase, including how and when they were actually administered, with as much detail as possible to facilitate attempts at replication | Yes |
| Procedural fidelity | 17 | Describe how procedural fidelity was evaluated in each phase | Yes |
| ANALYSIS |  |  |  |
| Analyses | 18 | Describe and justify all methods used to analyze data | Yes |
| **RESULTS** |  |  |  |
| Sequence completed | 19 | For each participant, report the sequence actually completed, including the number of trials for each session for each case. For participant/s who did not complete, state when they stopped and the reasons | Not applicable, as this manuscript reports a study protocol and no data have been collected yet. |
| Outcomes and estimation | 20 | For each participant, report results, including raw data, for each target behavior and other outcome/s | Not applicable, as this manuscript reports a study protocol and no data have been collected yet. |
| Adverse events | 21 | State whether or not any adverse events occurred for any participant and the phase in which they occurred | Not applicable, as this manuscript reports a study protocol and no data have been collected yet. |
| **DISCUSSION** |  |  |  |
| Interpretation | 22 | Summarize findings and interpret the results in the context of current evidence | Not applicable, as this manuscript reports a study protocol and no data have been collected yet. |
| Limitations | 23 | Discuss limitations, addressing sources of potential bias and imprecision | Yes |
| Applicability | 24 | Discuss applicability and implications of the study findings | Not applicable, as this manuscript reports a study protocol and no data have been collected yet. |
| **DOCUMENTATION** |  |  |  |
| Protocol | 25 | If available, state where a study protocol can be accessed | This study has been registered on clinicaltrial.gov (NCT06445140). |
| Funding | 26 | Identify source/s of funding and other support; describe the role of funders | This study is supported by the Ligue Nationale Contre le Cancer following a competitive peer-review process, under grant number 18195. |

**S2: EMA assessment items: The 5 versions of the questionnaire for TPB constructs, dietary intake and fatigue**

**Motivation toward physical activity**

**Version 1**

- Engaging in physical activity for at least 10 minutes in the next few hours could help me manage my stress.
- In my opinion, doing physical activity for at least 10 minutes in the next few hours would be enjoyable.
- For me, doing physical activity for at least 10 minutes in the next few hours would be approved by most of the important people in my life.
- I feel capable of being physically active for at least 10 minutes in the next few hours.
- I am going to engage in physical activity for at least 10 minutes in the next few hours.

**Version 2**

- For me, engaging in physical activity for at least 10 minutes in the next few hours would be beneficial for my health.
- In my opinion, doing physical activity for at least 10 minutes in the next few hours would be a source of pleasure.
- People close to me might encourage me to engage in physical activity for at least 10 minutes in the next few hours.
- It would be easy for me to do physical activity for at least 10 minutes in the next few hours.
- I intend to engage in physical activity for at least 10 minutes in the next few hours.

**Version 3**

- Doing physical activity for at least 10 minutes in the next few hours could help me manage my fatigue.
- For me, doing physical activity for at least 10 minutes in the next few hours could bring a sense of pleasure.
- My doctor would approve of me doing physical activity for at least 10 minutes in the next few hours.
- I think I can do physical activity for at least 10 minutes in the next few hours.
- I have planned to engage in physical activity for at least 10 minutes in the next few hours.

**Version 4**

- Engaging in physical activity for at least 10 minutes in the next few hours would be beneficial for my health.
- For me, doing physical activity for at least 10 minutes in the next few hours would be pleasant.
- In my opinion, my family members would want me to engage in physical activity for at least 10 minutes in the next few hours.
- Despite fatigue, I feel that I can do physical activity for at least 10 minutes in the next few hours.
- I plan to engage in physical activity for at least 10 minutes in the next few hours.

**Version 5**

- In my opinion, doing physical activity for at least 10 minutes in the next few hours would be good for my health.
- For me, engaging in physical activity for at least 10 minutes in the next few hours could be an enjoyable experience.
- People close to me would accept me engaging in physical activity for at least 10 minutes in the next few hours.
- I have the ability to do physical activity for at least 10 minutes in the next few hours.
- I estimate that my chances of doing physical activity for at least 10 minutes in the next few hours are high.

**Motivation toward nutrition**

**Version 1**

- Eating enough at my next meal would be beneficial for my health (>8/10 on the intake scale).
- In my opinion, my next meal will be enjoyable.
- Eating enough at my next meal would be approved by most of the important people in my life (>8/10 on the intake scale).
- I have the ability to eat enough at my next meal, even if I don't enjoy the taste (>8/10 on the intake scale).
- I intend to include enough at my next meal (>8/10 on the intake scale).

**Version 2**

- In my opinion, the quantity of my next meal is an important factor for my well-being.
- In my opinion, my next meal will be pleasant.
- People close to me would encourage me to eat enough at my next meal (>8/10 on the intake scale).
- I feel capable of eating enough at my next meal (>8/10 on the intake scale).
- I estimate that my chances of eating enough at my next meal are high (>8/10 on the intake scale).

**Version 3**

- Eating enough at my next meal is important to manage my fatigue (>8/10 on the intake scale).
- I will enjoy eating my next meal.
- My doctor would encourage me to eat enough at my next meal (>8/10 on the intake scale).
- It will be easy for me to eat enough at my next meal (>8/10 on the intake scale).
- I will eat enough at my next meal (>8/10 on the intake scale).

**Version 4**

- Eating enough at my next meal would be helpful to stay in good shape (>8/10 on the intake scale).
- Eating my next meal will be a source of pleasure for me.
- In my opinion, my family members would want me to eat enough at my next meal (>8/10 on the intake scale).
- I am capable of making the effort to eat enough at my next meal, even if I’m not hungry (>8/10 on the intake scale).
- I intend to eat enough at my next meal (>8/10 on the intake scale).

**Version 5**

- It is important that I eat enough at my next meal, even if I am not hungry (>8/10 on the intake scale).
- In my opinion, my next meal could bring me a sense of pleasure.
- People close to me would encourage me to eat enough at my next meal (>8/10 on the intake scale).
- I can eat enough at my next meal without difficulty (>8/10 on the intake scale).
- I plan to make the effort to eat enough at my next meal (>8/10 on the intake scale).

**Fatigue:** The patient answers the item: “At the moment, I feel tired” with a 7-point numeric scale with threshold values: 1 Not at all – 3 A bit – 5 Quite – 7 Very much.

**Dietary intake:**

Dietary intake will be assessed using a daily 10-point numeric rating scale based on the ingesta-VAS. The item asks: ‘If you consider that when you are in good health, you usually eat 10 out of 10, how much do you currently eat on a scale from 0 to 10?’

During the EMA, this item will be administered daily to assess participants’ current level of food intake: “If you consider that when you are healthy you eat at 10/10, today you ate at?”, through a 10-point numeric scale with: 1 I don’t eat anything – 10 I eat as usual.

- **S3: Visual representation of the display of the UniQ application**

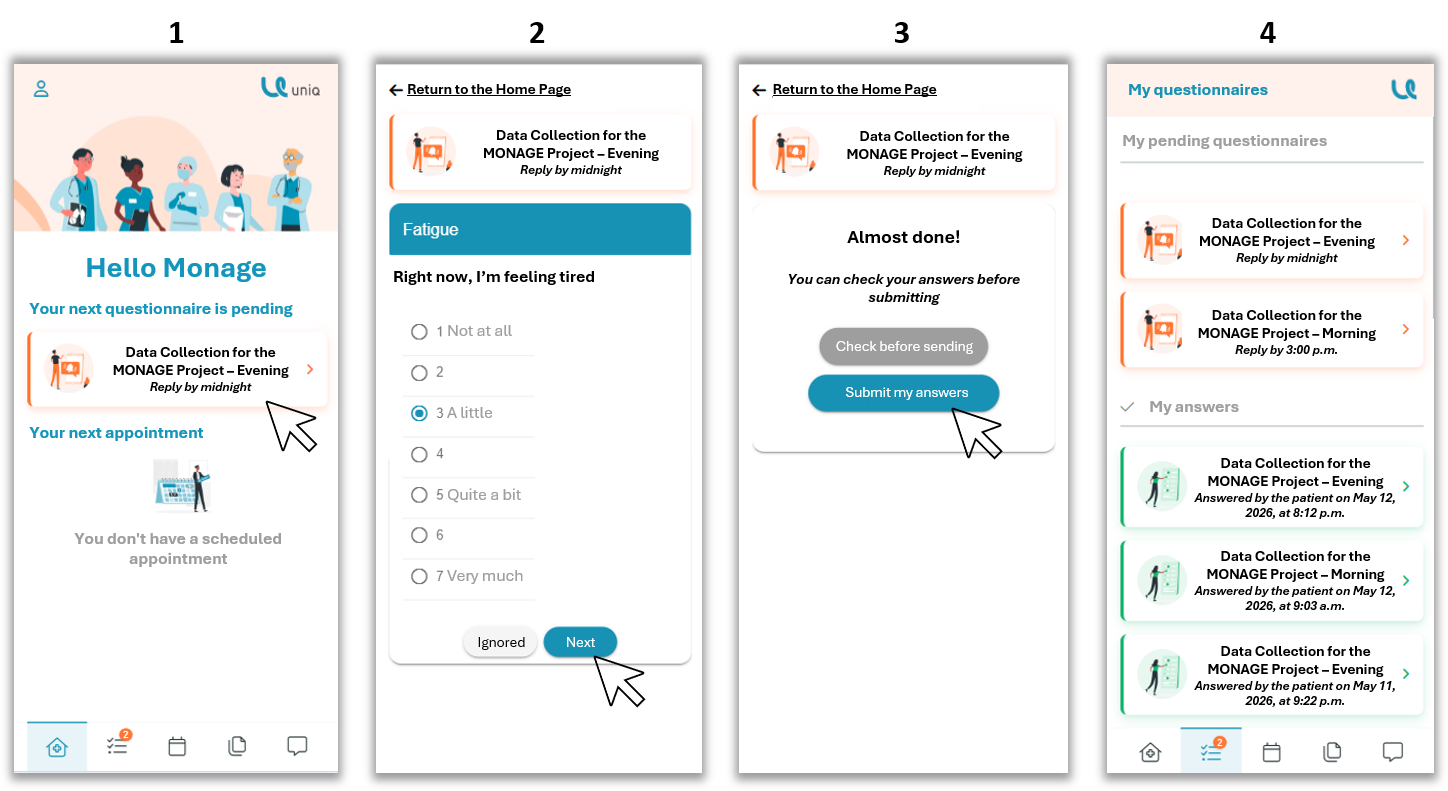
The patient can directly access the questionnaire available on the application interface (1). By clicking on it, they can answer the questions directly (2), submit the completed questionnaire (3), and finally access the remaining available questionnaires as well as their previous responses (4). The original content of the app is in French, but is translated into English below.

- **S4: Self-monitoring of behaviors conducted on the UniQ app**

During phase B of the intervention, participants completed a self-administered questionnaire every evening to report their physical activity and nutritional behaviours during the day. This questionnaire operationalized the Behaviour Change Technique (BCT) “self-monitoring of behaviour”, as participants were asked to record their daily behaviours and relevant characteristics of these behaviours.

- Physical activity:

Participants reported whether they had engaged in three types of physical activity during the day: (1) cardiorespiratory activities, such as walking or cycling; (2) muscle-strengthening activities; and (3) stretching and/or balance activities. For each type of activity performed, participants reported its duration using four categories: <15 min, 15–30 min, 30 min–1 h, or >1 h. They also rated the perceived intensity of each activity on a numeric rating scale from 1 to 10. Finally, participants reported the context in which the activity was performed: independently, accompanied by a relative, or supervised by a rehabilitation professional.

- Nutrition:

Participants rated their overall daily nutritional intake on a 10-point scale (1–10). They also reported the number of times they had consumed protein during the day (0, 1, 2, or >2 times) and whether they had consumed a snack. When a snack was reported, participants indicated whether it consisted of an oral nutritional supplement (ONS).

This daily questionnaire was completed each evening throughout phase B and was used to document participants’ day-to-day physical activity and nutritional behaviours.
